# Self-collection: an appropriate alternative during the SARS-CoV-2 pandemic

**DOI:** 10.1101/2020.04.09.20057901

**Authors:** Michael C. Wehrhahn, Jennifer Robson, Suzanne Brown, Evan Bursle, Shane Byrne, David New, Smathi Chong, James P. Newcombe, Terri Siversten, Narelle Hadlow

**Affiliations:** Douglass Hanly Moir Pathology, 14 Giffnock Ave Macquarie Park NSW 2113 Australia; Sullivan Nicolaides Pathology, 24 Hurworth St, Bowen Hills QLD 4006 Australia; Department of Endocrinology & Diabetes, Sir Charles Gairdner Hospital Hospital Ave, Nedlands, WA 6009 Australia; Clinipath Pathology 310 Selby St, North Osborne Park WA 6017, Australia

## Abstract

**BACKGROUND:** Swabs for SARS-CoV-2 are routinely collected by health care workers, putting them at risk of infection and requiring use of personal protective equipment (PPE). Self-collected swabs offer many advantages provided detection rate of SARS-CoV-2 and other respiratory viruses is not compromised.

**METHODS:** In a prospective study, patients attending dedicated COVID-19 collection clinics were offered the option to first self-collect (SC) nasal and throat swabs prior to health worker collection (HC). Two different laboratory services participated, with HC at Site 1 collecting nasal and throat swabs and at Site 2 nasopharyngeal (NP) and throat swabs. Samples were analysed for SARS-CoV-2 as well as common respiratory viruses. Concordance of results between methods was assessed using Cohen’s kappa (κ).

**RESULTS:** Of 236 patients sampled by HC and SC, 25 had COVID-19 (24 by HC and 25 by SC) and 63 had other respiratory viruses (56 by HC and 58 by SC). SC was highly concordant with HC (κ = 0.890) for all viruses including SARS-CoV-2 and more concordant than HC to positive results by any method (κ = 0.959 vs 0.933).

**CONCLUSIONS:** Self-collection of throat and nasal swabs offers a reliable alternative to health worker collection for the diagnosis of SARS-CoV-2 and other common respiratory viruses. High viral load of SARS-CoV-2 throughout the respiratory tract and sensitive molecular methods may explain these findings. Self-collection also provides patients with easier access to testing, reduces the exposure of the community and health workers to those undergoing testing and reduces the requirement for PPE.

## Introduction

On the 11^th^ March 2020, the World Health Organization (WHO) announced COVID-19 as a pandemic.^1^ The WHO Director-General issued a call for urgent action and encouraged all countries to ‘innovate and learn’ in their response to this crisis.

Demands on health services have increased and a commensurate decrease in availability of personal protective equipment (PPE) has occurred whilst the protection of health staff and the community remain paramount. Self-collected swabs in the community for SARS-CoV-2, the agent of COVID-19, and for other respiratory viruses offers potential significant benefit in the current pandemic by reducing requirement for PPE, and limiting exposure of patients and staff to infection.

Self-collection for respiratory viruses is not a new concept. Benefits include increased convenience and access for patients and timeliness of a sample receipt.^2,3^ Patients report self-collected nasal swabs are easy to perform^2,4,5^ and highly acceptable.^2,4^ A meta-analysis of 9 studies comparing self-collect (SC) and health care worker collect (HC) for influenza testing reported a pooled sensitivity of 87% and specificity of 99% for SC compared to HC^6^ however sensitivity for other respiratory viruses was not studied. Irving et al^7^ studied paired samples from 240 adults and found sensitivity using nasal or nasopharyngeal (NP) collection for influenza did not vary significantly when using a highly sensitive molecular test.^7^ A study in 230 children reported equivalent sensitivity for all respiratory viruses except respiratory syncytial virus (RSV) when comparing nasal swab and NP aspirate.^8^ Larios et al^9^ demonstrated that using flocked swabs and sensitive molecular methods, equivalent sensitivity and specificity was obtained for 76 matched self-collected mid-turbinate nasal swabs and NP swabs in 38 individuals for a range of respiratory viruses including human coronaviruses (hCoV 229E/NL63 and hCoV OC43/HKU1).

Recent reports on SARS-CoV-2 in respiratory specimens indicate early high viral loads in symptomatic and asymptomatic patients in a variety of clinical specimens including nasal and throat swabs, sputum and saliva samples.^10-14^ Wang et al reported that in 205 patients with COVID-19 the highest positive rates were found from bronchoalveolar lavage fluid, sputum and nasal swabs respectively.^15^ Wolfel^14^ and colleagues reported that in hospitalized cases of COVID-19 there was no discernible difference between NP and throat swabs with high viral load present in both specimens early in the illness and suggested that simple throat swabs may provide sufficient sensitivity when patients are first tested with mild symptoms of COVID-19.

The aim of this study was to compare prospectively the performance of HC with separate SC nasal (SCN) and throat swabs (SCT) and the combination of the two (SCNT) for respiratory viruses including SARS-CoV-2.

## Methods

This study was conducted across two laboratory sites (Site 1 and Site 2) and had ethics approval from the Western Australian branch of the Australian Medical Association, with all participants providing informed consent. For a period of one week in March 2020, patients presenting for SARS-CoV-2 testing at dedicated COVID-19 collection rooms were offered participation in the study. Demographic data was recorded including the address postcode to assess the Index of Education and Occupation (IEO) which assesses education level based on a scale of 1 to 5 with 5 being the highest level of education.^16^ A questionnaire assessing acceptability of SC based on that of Akmatov^4^ was provided to patients. Printed instructions including diagrams were provided on how to collect throat and nasal swab (See Supplementary Information). Self-collection kits included two swab packets each containing a single swab and screw-top container with 2mL liquid Amies medium, a tongue depressor and a zip lock sample bag. SC samples were taken immediately prior to trained HC samples to reduce ‘training bias.’ For SC and HC at Site 1 and SC at Site 2, open-cell polyurethane foam swabs (Σ Transwab^®^ ref MW940, Medical Wire & Equipment (MWE), Wiltshire, England) were used. Throat swabs were collected from the posterior throat and tonsil areas while nasal swabs were inserted as far as comfortably possible and at least 2-3 cm inside one nostril, rotating the swab 5 times and leaving in place for 5-10 seconds. For HC at Site 2, a flocked NP swab and a foam throat swab (Σ Transwab^®^ ref MW819 and MW940) were used. In addition, because the expected SARS-CoV-2 positivity rate at the time was estimated to be less than 1%, a subset of 24 patients recently diagnosed with COVID-19 performed SC in their homes.

At site 1, testing for SARS-CoV-2 was on the Allplex™ 2019-nCoV Assay (Seegene, Seoul, South Korea) and followed sample extraction using MagNA Pure 96 (Roche, Basel, Switzerland) with amplification utilising CFX96 Touch RT-PCR Detection Systems (BioRad, Hercules, California USA). Samples were confirmed as SARS-CoV-2 positive if all three gene targets (E/RdRp and N genes) were detected within 40 cycles. At site 2, the same extraction method was used. Testing for SARS-CoV-2 was performed using an in-house developed Taqman assay targeting the E gene.^17^ All positive samples then underwent 3 supplementary RT-PCRs targeting the N gene.^18^ Both laboratories utilised the Seegene RV Essential assay to detect other respiratory viruses (influenza A, influenza B, parainfluenza, RSV, human metapneumovirus (HMPV), adenovirus and rhinovirus).

### Statistical methods

A positive result on either HC or SC was defined as the benchmark result All Positives (AP). Concordance between HC and SC swabs and AP was calculated using Cohen’s Kappa (κ), which measures agreement between the categorical assignments given by two methods. The statistic takes values typically between zero and one. A κ >0.80 indicates very good agreement, while κ=1 indicates perfect concordance. Cycle threshold (Ct) values were recorded for all positive test results as a surrogate measure for viral load. Mean Ct was compared between HC and SCNT (combined category using the lowest Ct of either SCN or SCT), using linear mixed effects models, with a random effect for patient identification. HC and SC SARS-CoV-2 positivity rates were compared with Pearson’s χ^2^ test.

From power calculations assuming a significance level of 5% and a null hypothesis of low concordance between the HC and SC methods (H_0_: κ=0.3), there was at least 80% power to detect a concordance of 0.6 or more with a sample size of 66. Significance level α was set at 0.05, however for concordance and regression analyses, a Bonferroni multiple testing correction was applied such that minimum α’=0.05/8=0.0063. Statistical analyses were completed in the R statistical computing environment,^19^ including the package *irr*.

## Results

A total of 236 participants across the two sites took part in this study. Median age of participants was 40 (range 9-81) years and 60% were female. Twenty-five patients were positive for SARS-CoV-2 and 63 patients positive for other common respiratory viruses. For SARS-CoV-2 cases, 24/25 were detected by HC and 25/25 by SC. For common respiratory viruses 56/63 (89%) were detected by HC and 58/63 (92%) by SC (Table 1). A positive result on either HC or SCNT was included in the group AP.

**Table 1:** Summary of COVID19 cases, other respiratory cases and negative test results from both sites, with corresponding detections under the HC and SCNT methods.

| N=236 | Test Result | Site 1 | Site 2 | All Patients |
| --- | --- | --- | --- | --- |
| **HC** | Negative | 38 | 117 | 155 (65.7%) |
|  | Other Respiratory | 20 | 36 | 56 (23.7%) |
|  | COVID19 | 12 | 12 | 24 (10.2%) |
| **SCNT** | Negative | 35 | 118 | 153 (64.8%) |
|  | Other Respiratory | 23 | 35 | 58 (24.6%) |
|  | COVID19 | 12 | 13 | 25 (10.6%) |
| **AP** | Other Respiratory | 23 | 40 | **63** (26.7%) |
|  | COVID19 | 12 | 13 | **25** (10.6%) |

HC: Health worker Collect; SCNT: Self Collect Nasal and Throat; AP: All Positives (positive results from either HC or SCNT).

Table 2 summarises the respiratory viruses detected by the different methods of collection. At Site 1, co-detection of rhinovirus (Ct 29) + influenza A (Ct 41) was found in one patient by SC only and RSV (Ct 24) + rhinovirus (Ct 35) in one patient by HC only. Two Parainfluenza cases and one rhinovirus case were detected only by SC. Overall the detection rate was 6% higher in SC compared with HC swabs for non-SARS-CoV-2 respiratory viruses which equated to 3/20 (15%) additional positive results. At Site 2, no co-detections occurred. Collection of samples for the 13 SARS-CoV-2 positive patients ranged from 2 to 9 days following onset of symptoms with a mean of 4.8 days. One positive patient retested 6 days after symptom onset using the screening E-gene assay, was detected only on SCN but not the HC. A second positive patient was detected using HC and SCT but not SCN. Of the patients with detectable respiratory viruses other than SARS-CoV-2, at site 1, 8/23 (35%) had virus only detectable on one of SCN or SCT while the proportion was 14/35 (40%) at site 2.

**Table 2:** Summary of COVID-19 and other respiratory illnesses detected under the HC, SCN, SCT, SCNT methods and positives from all methods (AP), at the two collection sites.

| **Site 1** | **HC** | **SCN** | **SCT** | **SCNT** | **AP** |
| --- | --- | --- | --- | --- | --- |
| Rhinovirus | 15 | 15 | 14 | 16 | 16 (22.9%) |
| Influenza B | 2 | 1 | 2 | 2 | 2 (2.9%) |
| RSV | 1 | 1 | 1 | 1 | 1 (1.4%) |
| Adenovirus | 1 | 0 | 1 | 1 | 1 (1.4%) |
| Parainfluenza | 0 | 2 | 1 | 2 | 2 (2.9%) |
| HMPV | 1 | 1 | 0 | 1 | 1 (1.4%) |
| **Total Other Respiratory** | **20 (28.6%)** | **20** | **19** | **23 (32.9%)** | **23 (32.9%)** |
| **SARS-CoV-2 (E,N,RdRp gene)** | **12 (17.1%)** | **5/5\*** | **5/5\*** | **12 (17.1%)** | **12 (17.1%)** |
| Total undergoing HC and SC | 70 (100%) |  |  | 70 (100%) | 70 (100%) |

| **Site 2** | **HC** | **SCN** | **SCT** | **SCNT** | **AP** |
| --- | --- | --- | --- | --- | --- |
| Rhinovirus | 23 | 19 | 17 | 22 | 25 (15.1%) |
| Influenza B | 1 | 1 | 0 | 1 | 1 (0.6%) |
| RSV | 1 | 1 | 1 | 1 | 1 (0.6%) |
| Adenovirus | 2 | 2 | 1 | 3 | 4 (2.4%) |
| Parainfluenza | 7 | 4 | 6 | 6 | 7 (4.2%) |
| HMPV | 2 | 2 | 2 | 2 | 2 (1.2%) |
| **Total Other Respiratory** | **36 (28.6%)** | **29** | **27** | **35 (21.1%)** | **40 (24.1%)** |
| **SARS-CoV-2 (E gene\*\*)** | **12 (7.2%)** | **12** | **11** | **13 (7.8%)** | **13 (7.8%)** |
| Total undergoing HC and SC | 166 (100%) | 166 | 166 | 166 (100%) | 166 (100%) |

HC: Health worker Collect; SCN: Self Collect Nasal; SCT: Self Collect Throat; SCNT: Self Collect Nasal and Throat; AP: All Positives (positive results from either HC or SCNT); RSV: Respiratory Syncytial Virus; HMPV: human metapneumovirus. \*only a subset of 5 patients at Site 1 had nasal and throat swabs tested individually. \*\*All patients had supplementary N gene testing: HC 13; SCN 13; SCT 11; SCNT 13 detected.

When all detections by HC and SCNT were compared with AP, the sensitivity of SCNT and HC to detect COVID-19 was 1.0 (95%CI: 0.86-1) and 0.96 (95%CI: 0.8-1) respectively; for other respiratory viruses it was 0.94 (95%CI: 0.87-0.98) and 0.91 (95%CI: 0.83-0.96) respectively.

Table 3 summarises concordance between AP and each collection method. Both SCNT and HC showed very high concordance with AP at each site and overall, with SCNT slightly higher (κ=1, 0.934, 0.959 at Site1, Site2, Combined Sites) than HC (κ=0.929, 0.934, 0.933). Additionally, SCNT was highly concordant with HC (κ=0.929, 0.863, 0.890 at Site 1, Site 2, Combined Sites). When Ct values for COVID-19 cases were compared by collection method (Figure 1), mean E-gene Ct did not differ between HC and SCNT or SCN (p=0.236, 0.083, against α’=0.0083) but was significantly higher in SCT compared with HC (β=7.31, p<0.001). Mean N-gene Ct was not significantly higher in SCNT compared with HC (p=0.041; α’=0.0083) but was higher in SCN and SCT (β=4.00, p=0.006; β=7.63, p<0.001). In rhinovirus cases (Figure 2), mean Ct was not significantly higher in SCNT compared with HC (p=0.036; α’=0.017) but was higher in SCN and SCT (β=2.50, p=0.002; β=6.68, p<0.001). In Parainfluenza cases, mean Ct differed between HC and SCN (β=4.67, p=0.014) but not the other methods (SCNT v HC, p=0.231; SCT v HC, p=0.119; α’=0.017).

**Table 3:** Concordance (Cohen’s κ) between (i) AP and HC, SCN, SCT and SCNT; and (ii) HC and SCNT. A value of 1 indicates the method detected all COVID-19 and other respiratory cases, while a value above 0.9 indicates a very high level of detection of all respiratory cases (AP).

| Concordance with AP | HC | SCN | SCT | SCNT |
| --- | --- | --- | --- | --- |
| Site 1 | 0.929 | 0.905<sup>*</sup> | 0.872<sup>*</sup> | 1 |
| Site 2 | 0.934 | 0.835 | 0.789 | 0.934 |
| Combined Sites | **0.933** | 0.858 | 0.817 | **0.959** |
| Concordance between HC and SCNT | Site 1 |  | Site 2 | Combined Sites |
|  | **0.929** |  | **0.863** | **0.890** |

HC: Health worker Collect; SCN: Self Collect Nasal; SCT: Self Collect Throat; SCNT: Self Collect Nasal and Throat; AP: All Positives (positive results from either HC or SCNT). P-value <0.001 for each concordance test. \* SCN and SCT concordance on reduced set of individuals for Site 1 (only 5 of 12 SARS-CoV-2 patients had SCN and SCT testing individually performed).

**Figure 1:**
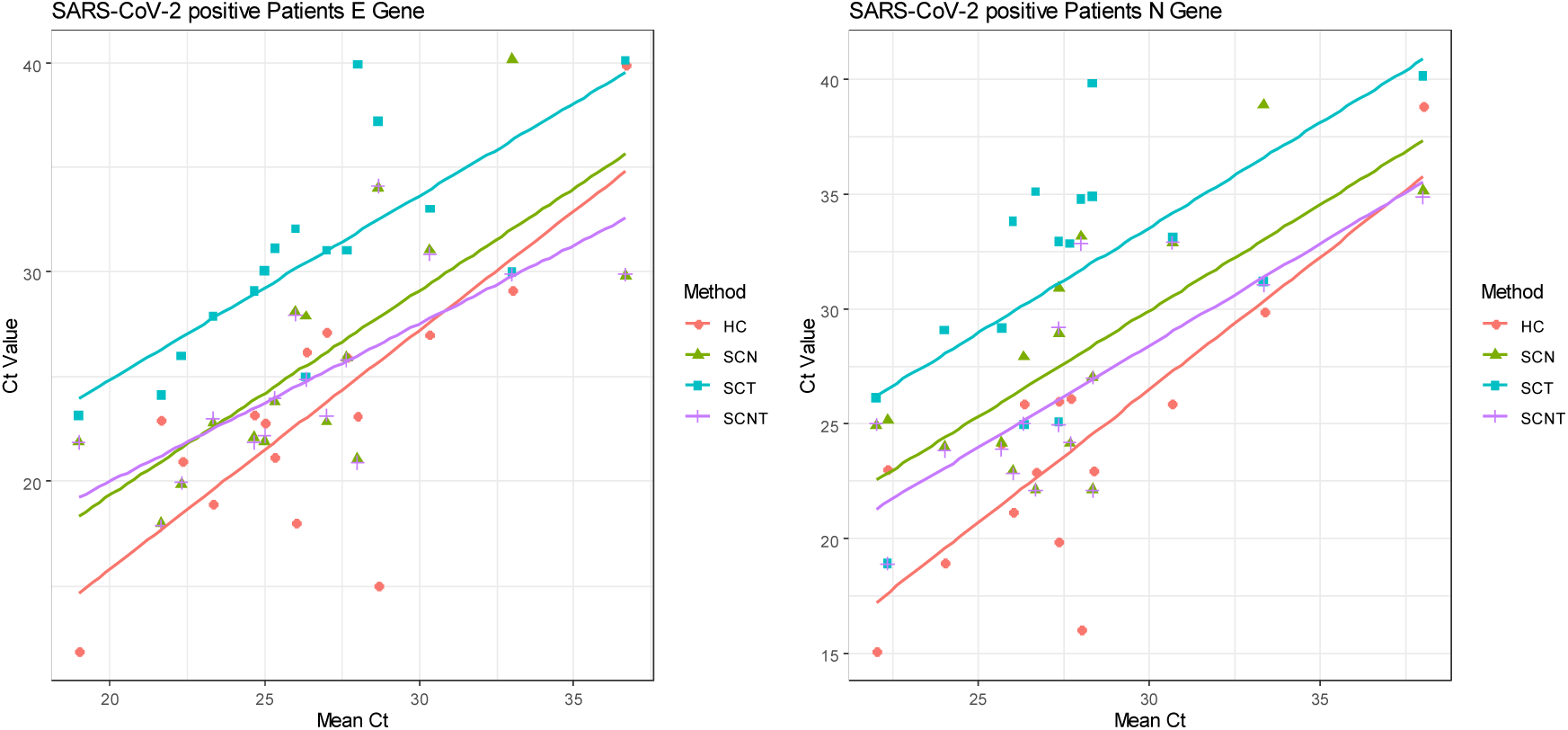
E-gene and N-gene Ct values obtained by the different collection methods for SARS-CoV-2 positive patients at both sites.

**Figure 2:**
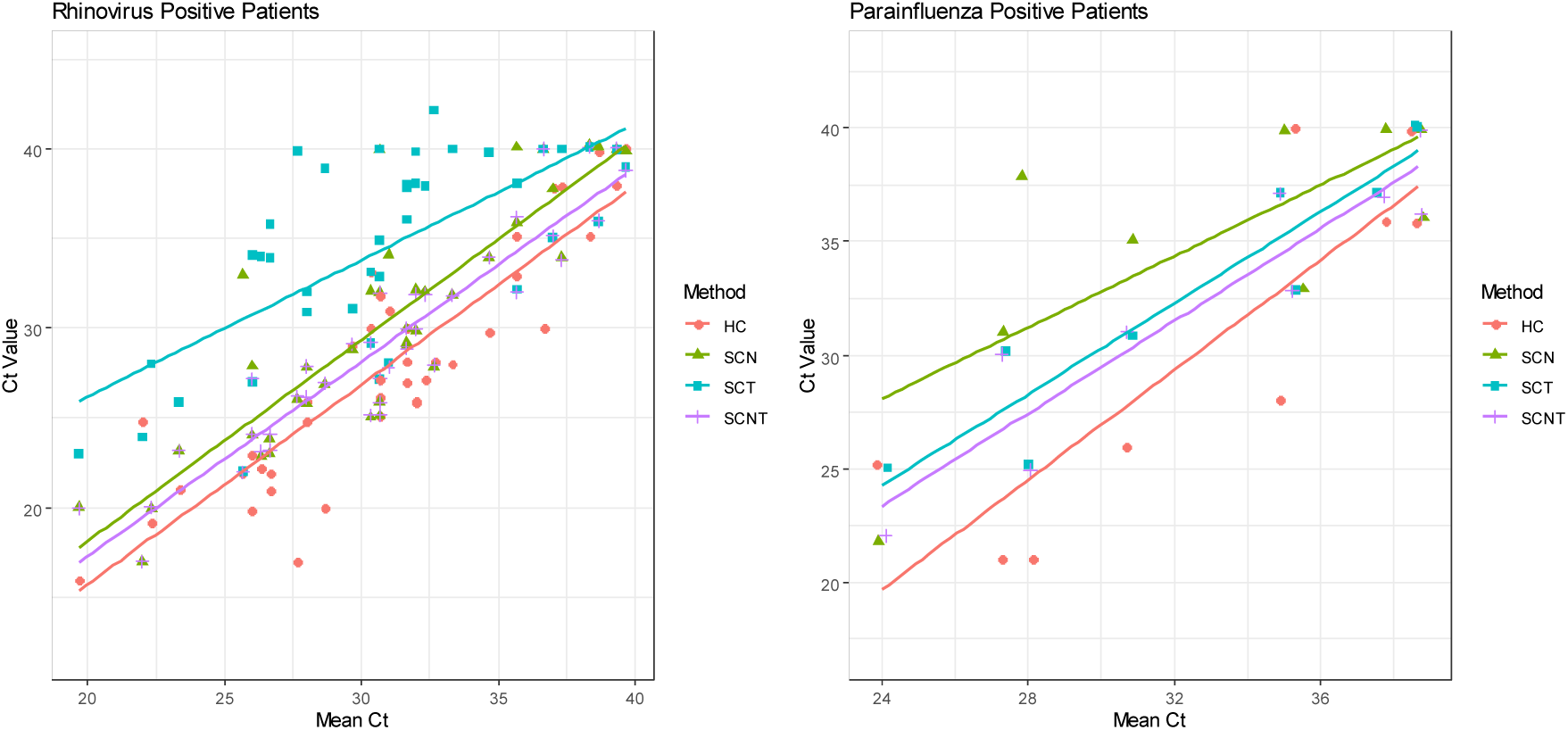
Ct values obtained by the different collection methods for rhinovirus and Parainfluenza positive patients at both sites.

At Site 1 an analysis of acceptability was performed using a questionnaire and was completed by 42/70 (60%) participants with 31/42 (74%) preferring self-collection over trained collectors, with all considering it acceptable. Analysis of the IEO found that the Median (LQ, UQ) IEO was 3 (2, 4) with participants identified across all educational levels but the majority (30/42, 71%) were in the 3 lowest education levels and a smaller proportion (12/42, 29%) in the highest 2 levels.

Following this study, Site 1 has since processed a small percentage of SC swabs (7% of all collections). There was no significant difference in the SARS-CoV-2 detections between HC with 242/13851 (1.8%) and SC with 20/1035 (1.9%) (p=0.753 from χ^2^ test).

## Discussion

In our group of 236 ambulatory, literate, mostly adult patients, the performance of self-collected nasal and throat swabs was at least equivalent to that of health care worker collected swabs for the detection of SARS-CoV-2 and other respiratory viruses.

This study included two different sites using two different methods of HC (combined N + T and combined NP + T) and also employed two different molecular strategies for detection of SARS-CoV-2. As such these findings are more widely applicable.

At Site 1 where SCNT was compared with HC using the same swab and collection methods, for the 12 patients testing positive to SARS-CoV-2 there was complete concordance between HC and SC samples even though on average 2.5 days had lapsed. In the remaining SARS-CoV-2 negative patients, SC detected 3 additional respiratory viruses, with the overall positivity rate increasing from 34% to 40%. However, the additional 3 SC detections were weak positives based on high Ct values (33-40).

At site 2 where comparative HC involving a NP and T swab occurred at the same time as the SCN and SCT for the SARS-CoV-2 positive patients, SCNT detected all 13 positive patients while one patient was negative by HC. Detection of other respiratory viruses by SCNT was highly concordant with HC detecting only 1 less respiratory virus and may relate to the fact that SCNT sampling was compared with NP +T sampling.

When data from each site was combined, concordance between SCNT or HC with the All Positive rate was very high, slightly favouring SCNT. The similar SARS-CoV-2 percent-positivity rate in ongoing comparison data between those having only HC or SC provides further reassurance that SCNT is equivalent to HC.

The advantages of self-collection are evident and even more important at a time of global health crisis. Self-collection greatly reduces the number of patients requiring trained health worker collection and PPE, thus preserving the limited supplies of PPE. Access to testing is increased, as swab kits can be provided quickly by clinicians or available at dedicated COVID-19 collection centres aiding timeliness of testing^2,3^ which is critical in the current pandemic. There is increased safety for both patients and staff using a SC model as exposure to others is limited.

Further, data from patients at site 1 suggests that SC is accessible and achievable over a range of education levels with all finding SC acceptable and the majority having a preference for this method over HC as has previously been reported.^2,4,5^ This may relate to the ability of patients to control the comfort level of throat and nasal collection better than a trained collector can.

We chose to trial SCN and SCT swabs rather than NP collections because the latter is technically more difficult and uncomfortable for patients. Literature suggests that collection of mid-turbinate nasal swabs is comparable in performance to collection of NP swabs for respiratory viruses including other coronaviruses.^9^ We chose to perform nasal swabs given that mid-turbinate swabs with a safety stopping point are generally not as widely used and more uncomfortable than nasal swabs.

Recent studies suggest there is a high viral load in patients with early COVID-19 across the upper and lower respiratory tracts, including nasal and throat sites^10-12,14^ as well as in saliva,^13^ even in asymptomatic, mild or prodromal states. Wolfel et al^14^ noted no discernible difference between nasopharyngeal and oropharyngeal viral loads and detection rates in hospitalized cases of COVID-19 and noted that simple throat swabs provide sufficient sensitivity in early infections. Given these high viral loads throughout the respiratory tract it may be that requiring NP sampling is not as significant for SARS-CoV-2 as for some other respiratory viruses. It may also be that sensitive and specific PCR methods for viral detection are improving the sensitivity of a range of sample and collection methods as shown for a range of respiratory viruses but also Group A Streptococcal detection.^9,10^ We hypothesize that the high viral load of SARS-CoV-2 and sensitive molecular techniques may explain the equivalent sensitivity of SC to HC samples in COVID-19 patients. Additionally viral load at different sites may differ with disease evolution and the SARS-CoV-2 positive patients in this study were tested over a range of 2 to 9 days from symptom onset.

Our data support the decision by the Communicable Disease Network of Australia (CDNA)^21^ to recommend sampling of both nasal and throat sites for the diagnosis of respiratory viruses including for SARS-CoV-2, due to the concern of a possible missed diagnosis if only one site is sampled. This was the case for two COVID-19 positive patients on SC who were only diagnosed by SCN and another only by SCT. If only one swab site was obtainable, our data suggests the nasal may be the better swab site for the diagnosis of COVID-19 as it had greater concordance with the AP group and showed consistently lower Ct values in the order of 100-1000 fold higher viral load (data not shown).

Limitations of this study include the limited number of positive SARS-CoV-2 patients and modest number of other positive respiratory virus cases with the exception of rhinovirus. Further data on self-collection would be helpful to confirm these findings. In the setting of limited resources, both in terms of PPE and health care workers, these findings may be important for other health services. Furthermore, we have instituted use of a single swab to sample both throat then nasal sites. This has the potential to preserve limited supplies of swabs and also provide additional efficiencies in the laboratory as only preparation of a single sample per patient is required.

## Conclusion

The world is facing unprecedented demands on health care services and health resources during the COVID-19 pandemic. Innovative ways to address this crisis are required and we believe that this study provides early evidence that self-collection of throat and nasal swabs for SARS-CoV-2 offers an acceptable and reliable alternative to health care worker collected samples. This is achieved whilst preserving critically needed PPE supplies, optimizing the time to testing and reducing exposure of health care workers to potentially infected patients.

## Data Availability

Deidentified data may be made available on request

## Acknowledgement

We thank the Training and Patient Services departments, the Collection and Clinical Area Managers, Clinical Supervisors, Collection staff, and Molecular Laboratory staff without whom this study would not have been possible.

